# EXCLUSIVE BREASTFEEDING AMONG WORKING MOTHERS IN GHANA: EVIDENCE FROM THE 2022 GHANA DEMOGRAPHIC AND HEALTH SURVEY

**DOI:** 10.64898/2026.08.26.26361421

**Authors:** Abubakar Iddrisu Siddiq, Ishmael Saafu, Edmund Teye Borkor, Emmanuel Vondee, Felicia Tenpoka Sampana, Bernice Okine

**Author notes:** ***Corresponding author:*** *Abubakar Iddrisu Siddiq,.

## Abstract

**Background:** Exclusive breastfeeding may protect infants against common infections and support healthy growth and development. Working mothers may face constraints on exclusive breastfeeding arising from work schedules, separation from their infants, and inadequate breastfeeding support. National evidence on the individual, healthcare-related, and contextual factors associated with exclusive breastfeeding among working Ghanaian mothers appears to remain limited.

**Design:** Cross-sectional secondary analysis.

**Setting:** Nationally representative survey covering urban and rural communities across all 16 administrative regions of Ghana.

**Participants:** The analysis included 620 currently working mothers whose youngest living infants were aged 0–5 completed months and lived with them. The complete-case multivariable analysis included 619 mother–infant pairs.

**Primary outcome measure:** Current exclusive breastfeeding, defined using the standard 24-hour infant-feeding indicator. Infants were classified as exclusively breastfed when they received breast milk without water, formula, animal milk, other liquids, or solid or semi-solid foods during the preceding day or night. Oral rehydration solution, vitamins, minerals and prescribed medicines were permitted.

**Aim:** To estimate the prevalence of exclusive breastfeeding and examine its individual, healthcare-related and contextual correlates among working mothers of infants aged 0–5 months in Ghana.

**Methods:** Birth Recode data from the 2022 Ghana Demographic and Health Survey were analysed. Unweighted frequencies and survey-weighted percentages described the study population. Design-adjusted Wald tests assessed bivariate associations. Survey-weighted binary logistic regression estimated adjusted odds ratios (AORs) and 95% confidence intervals (CIs), accounting for sampling weights, primary sampling units, and strata.

**Results:** The survey-weighted prevalence of exclusive breastfeeding was 54.3% (95% CI: 49.2–59.3). Ethnicity, mode of delivery, region, and community poverty appeared to be statistically significant in the bivariate analyses. In the adjusted model, region was jointly associated with exclusive breastfeeding (p = 0.004). Mothers in the Northern (AOR = 4.93; 95% CI: 1.50–16.17) and Savannah (AOR = 4.22; 95% CI: 1.08–16.41) regions had higher odds than mothers in the Western Region. Mothers in low-education communities had lower odds than those in high-education communities (AOR = 0.54; 95% CI: 0.30–0.98). Although Guan mothers had higher odds than Akan mothers, the overall association with ethnicity was non-significant, and the estimate appeared imprecise. Maternal age, individual education, religion, parity, wealth, infant sex, antenatal care, postnatal care, and residence were not independently associated with exclusive breastfeeding.

**Conclusion:** The prevalence estimate suggests that slightly more than half of working mothers exclusively breastfed their infants. Regional and community differences appeared more pronounced than those associated with most measured individual characteristics. Regionally responsive breastfeeding support and practical community education may contribute to improved coverage. Workplace recommendations require further evidence because employment conditions were not measured directly.

**Strengths and limitations:**

□ The study used nationally representative data covering all 16 regions of Ghana and applied the survey weights, clusters and strata, and may thereby improve the generalisability of the estimates to working mothers of infants aged 0–5 months.
□ Exclusive breastfeeding was reconstructed using the complete standard 24-hour Demographic and Health Survey (DHS) feeding indicator, and mothers who had discontinued breastfeeding remained in the denominator, which may reduce the risk of overestimating prevalence.
□ The cross-sectional design did not establish temporal order or causality, while the 24-hour feeding measure may not reflect feeding practices throughout the period from birth to the interview.
□ Employment was measured only as current working status. Information on occupation, maternity leave, working hours, breastfeeding breaks, childcare arrangements and facilities for expressing and storing milk was unavailable.
□ Small sample sizes in some ethnic and regional categories may have produced wide confidence intervals. Community poverty and education were derived contextual proxies and may not have directly measured community resources, breastfeeding knowledge, or service availability.

## 1. Introduction

Exclusive breastfeeding may be an effective intervention for supporting infant survival, growth, and development [1]. The World Health Organization (WHO) recommends initiating breastfeeding within the first hour after birth and feeding infants exclusively with breast milk during their first six months. Safe and nutritionally adequate complementary foods should then be introduced while breastfeeding continues for two years or longer [2].

Exclusive breastfeeding is defined as feeding an infant only breast milk, including expressed breast milk, without water, formula, animal milk, other liquids or solid and semi-solid foods. Oral rehydration solution, vitamin and mineral supplements, and prescribed medicines are permitted under this definition. Breast milk may provide the energy and nutrients required during early infancy and contains antibodies and other immunological components that may protect infants against common childhood infections. Exclusive breastfeeding is associated with lower risks of diarrhoeal disease, acute respiratory infections, and infant mortality [3],[4].

Although the most immediate benefits of breastfeeding occur during infancy, its longer-term effects may extend beyond this period. Breastfed children may experience reduced risks of overweight, obesity, and diabetes later in life, while breastfeeding may reduce mothers’ long-term risks of breast and ovarian cancers [5]. Breastfeeding can also reduce household expenditure on breast-milk substitutes and the costs associated with treating preventable childhood illnesses [6]. These benefits appear to have contributed to the positioning of breastfeeding promotion as an important component of maternal, newborn and child health programmes [7].

Although exclusive breastfeeding has increased globally, its coverage appears to remain below international targets [8]. Approximately 48% of infants younger than six months were exclusively breastfed globally, and in 2024 the WHO recommended increasing this prevalence to at least 60% by 2030 [9]. Further progress may depend on addressing the social, economic and institutional conditions that affect mothers’ ability to initiate and sustain exclusive breastfeeding.

Ghana has implemented interventions such as breastfeeding counselling during antenatal and postnatal care, the Baby-Friendly Hospital Initiative and community-based infant and young child feeding education [10]. Recent studies indicate that approximately half of infants aged 0–5 months in Ghana were exclusively breastfed [11]. This national estimate may conceal differences between population groups and geographical areas. Breastfeeding practices can be influenced by maternal age, education, parity, ethnicity, household economic position, mode of delivery, healthcare use and community conditions [11],[11].

Working mothers may encounter particular difficulties in sustaining exclusive breastfeeding. Employment may require mothers to spend extended periods away from their infants, return to work before the infant reaches six months, or combine breastfeeding with demanding working schedules [12]. Short maternity leave, inadequate breastfeeding breaks, lack of childcare near the workplace and the absence of private facilities for expressing and storing breast milk may encourage the early introduction of formula, water or complementary foods. These conditions may affect both women in formal employment and those working in informal occupations [13],[14].

The effects of employment may not be uniform. A mother’s ability to continue exclusive breastfeeding may depend on her occupation, working hours, household resources, family support and access to breastfeeding information [11],[15]. Healthcare contacts during pregnancy and after childbirth may offer counselling and practical assistance, while regional and community conditions may determine the availability of breastfeeding support. Consequently, exclusive breastfeeding among working mothers may warrant examination within individual, healthcare-related, and contextual settings.

Previous Ghanaian studies have examined exclusive breastfeeding in the general population or within specific health facilities and occupations. National evidence focusing specifically on working mothers appears limited [15]. It is also unclear whether individual characteristics, maternal healthcare use, or community conditions may account for differences in exclusive breastfeeding within this population.

This study therefore assessed the prevalence and factors associated with exclusive breastfeeding among working mothers of infants aged 0–5 months in Ghana using data from the 2022 GDHS. The specific objectives were to examine the associations of individual-level, healthcare-related and contextual factors with exclusive breastfeeding. The study focused on associations and did not seek to establish causal effects. Its findings may support the development of breastfeeding interventions that respond to the circumstances of working mothers and the communities in which they live.

## 2. Design and Methods

### 2.1 Study Design and Data Source

A quantitative, cross-sectional analytical study was undertaken using secondary data from the 2022 Ghana Demographic and Health Survey. The 2022 GDHS constituted the seventh round of Ghana’s Demographic and Health Surveys [16]. Data collection took place from 17 October 2022 to 14 January 2023.

The survey collected nationally representative information on demographic characteristics, reproductive health, maternal and child health, nutrition, and infant-feeding practices, and covered urban and rural communities in all 16 administrative regions of Ghana [17]. The present analysis used the Birth Recode dataset because it contained information on maternal characteristics, childbirth, healthcare use and feeding practices for children born to interviewed women.

### Sampling design of the 2022 GDHS

The 2022 GDHS employed a stratified two-stage cluster sampling design. The sampling frame was based on enumeration areas created for the 2021 Ghana Population and Housing Census. Urban and rural areas within each of Ghana’s 16 regions were treated as separate sampling strata [17].

In the first stage, 618 clusters were selected using probability proportional to size. In the second stage, households were selected systematically from updated household listings within each cluster. Approximately 18,450 households were selected. Interviews were successfully completed in 17,933 households, representing a household response rate of approximately 99%. Of the 15,317 eligible women aged 15–49 years identified in the interviewed households, 15,014 completed the individual interview, producing a response rate of approximately 98% [16].

### Study population and eligibility criteria

The study population comprised currently working mothers with living infants aged zero to five completed months at the time of the survey. A mother was classified as currently working when she answered affirmatively to the DHS question concerning whether she was working at the time of the interview.

The Birth Recode dataset contained 34,663 birth records. Following restriction to each respondent’s most recent birth, eligible records were limited to children who were alive, aged zero to five completed months, and living with the interviewed mother. Finally, mothers who reported that they were currently working were retained.

This selection procedure yielded 620 eligible mother–infant pairs for the prevalence and descriptive analyses. One mother had an indeterminate response for the number of antenatal care visits. The complete-case multivariable analysis therefore included 619 mother–infant pairs.

The study population is described as working mothers of infants aged 0–5 months, rather than working lactating mothers. Mothers who had discontinued breastfeeding were retained because excluding them could remove non-breastfeeding mothers from the denominator and thereby potentially overestimate the prevalence of exclusive breastfeeding.

### Data collection instrument

The 2022 Ghana Demographic and Health Survey (GDHS) used standardised household, women’s, men’s, and biomarker questionnaires adapted to the Ghanaian context. Trained interviewers administered the questionnaires through face-to-face interviews using computer-assisted personal interviewing [16].

The women’s questionnaire collected information on respondents’ sociodemographic characteristics, reproductive histories, employment, antenatal care, delivery, postnatal care and child-feeding practices. Infant-feeding questions asked mothers to report all liquids, foods and supplements consumed by the child during the day and night preceding the interview. The use of a standard 24-hour reference period was consistent with the DHS approach to measuring current infant-feeding practices [18].

### Study variables Outcome variable

The outcome was current exclusive breastfeeding, measured using the standard 24-hour infant-feeding definition [18]. An infant was classified as exclusively breastfed when the mother reported that the child was currently receiving breast milk and had not consumed water, fruit juice, tea, coffee, formula, animal milk, other liquids, or solid or semi-solid foods during the preceding day or night.

The definition encompassed oral rehydration solution, vitamins, minerals, and prescribed medicines [19]. Exclusive breastfeeding was coded as 1, while non-exclusive breastfeeding was coded as 0.

### Individual-level variables

Individual-level variables comprised maternal age at delivery, educational attainment, religion, parity, ethnicity, household wealth, mode of delivery, and infant sex.

Maternal age at delivery was derived from the mother’s and child’s century month codes; it was subsequently categorised as younger than 20 years, 20–34 years, or at least 35 years. Education was classified as no education, primary education, and secondary or higher education. Religion was grouped as Christianity, Islam, and traditional religion or other.

Parity was categorised as primiparous for one birth, multiparous for two to four births and grand multiparous for at least five births. Ethnicity was classified as Akan, Ga/Dangme, Ewe, Guan, Mole-Dagbani, Grusi, Gurma, Mande, and other.

The original DHS wealth quintiles were recoded into three categories: the poorest and poorer quintiles formed the poor category, the middle quintile remained the middle category, and the richer and richest quintiles formed the rich category. Mode of delivery was classified as vaginal or caesarean, while infant sex was classified as male or female.

### Healthcare-related variables

The healthcare-related variables were antenatal care attendance and receipt of postnatal care. Antenatal care was classified as 0–3 visits and at least four visits. Postnatal care was coded as yes when the mother reported that the child received a health check following delivery or discharge and no when no such check was reported.

### Contextual variables

The contextual variables were urban or rural residence, region of residence, community poverty, and community education. Region was analysed using Ghana’s 16 administrative regions.

Community poverty was constructed at the survey-cluster level. For each cluster, the survey-weighted proportion of unique mothers in the Birth Recode dataset who belonged to the poorest or poorer wealth quintiles was calculated. The weighted national median of the cluster-level proportions was 23.5%. Clusters above this median were classified as high-poverty communities, while clusters at or below the median were classified as low-poverty communities.

Community education was operationalised as the survey-weighted proportion of unique mothers in each cluster who had no education or only primary education. The weighted national median was 33.3%. Clusters above the median were classified as low-education communities, while those at or below the median were classified as high-education communities.

These community indicators were intended to characterise the socioeconomic composition of the clusters represented in the Birth Recode dataset. They were not direct measurements of community infrastructure, services, or collective attitudes.

### Data management and statistical analysis

The dataset was cleaned, recoded, and analysed using Python 3. The Birth Recode variables were examined against their DHS variable labels before the study population and exclusive-breastfeeding indicator were constructed.

The women’s individual sampling weight was divided by 1,000,000 before application. All weighted analyses accounted for the primary sampling units and sampling strata supplied with the 2022 GDHS dataset. Unweighted frequencies and survey-weighted percentages were used to characterise the study population. The prevalence of exclusive breastfeeding was reported with a design-adjusted 95% confidence interval.

Design-adjusted Wald tests were used to assess the bivariate association between each independent variable and exclusive breastfeeding. Pearson chi-square tests that ignored the sampling design were not used because they may underestimate standard errors and produce misleading significance tests [20].

Survey-weighted binary logistic regression was used to examine factors independently associated with exclusive breastfeeding. Maternal age, education, religion, parity, ethnicity, household wealth, mode of delivery, infant sex, antenatal care, postnatal care, residence, region, community poverty and community education were entered into the adjusted model. Results were reported as adjusted odds ratios with 95% confidence intervals and p-values.

The logistic regression model was specified as:

where was the probability that a mother exclusively breastfed her infant, was the intercept, to were the regression coefficients, and to represented the individual, healthcare-related and contextual variables.

The overall adjusted model was assessed using a design-adjusted Wald test. A pseudo-Nagelkerke measure and survey-weighted classification accuracy were also calculated as descriptive measures of model performance. Statistical significance was set at a two-sided p-value below 0.05.

### Ethical considerations

The original 2022 Ghana Demographic and Health Survey (GDHS) protocol received ethical approval from the Ghana Health Service Ethics Review Committee and the ICF Institutional Review Board. Participants provided informed consent before the original interviews. The dataset used for this secondary analysis was anonymised and contained no direct personal identifiers. [21]

Authorisation to access and use the data was obtained from the DHS Programme. The present analysis involved no direct contact with participants, and no additional participant consent was required. [22]

### Methodological limitations

Although the cross-sectional design may identify factors associated with exclusive breastfeeding, it may not permit temporal or causal conclusions. The study identified associations and should not be interpreted as demonstrating that regional, ethnic, or community characteristics caused the observed breastfeeding differences.

The outcome was based on foods and liquids consumed during the preceding 24 hours. This method may primarily capture current feeding status and may classify an infant as exclusively breastfed even if the infant received other foods or liquids outside the reference period. [18] Maternal reports may also have been affected by recall bias. [18]

The analysis captured mothers’ current employment status but did not provide detailed measures of occupational characteristics, maternity leave, working hours, breastfeeding breaks, childcare arrangements or access to facilities for expressing and storing breast milk. The modest sample may also have produced small numbers within some ethnic and regional categories, potentially resulting in wide confidence intervals. In particular, the estimates for Guan ethnicity and several regions should be interpreted cautiously.

The community poverty and education variables were derived from the socioeconomic composition of survey clusters represented in the Birth Recode dataset. They may therefore be more appropriately interpreted as contextual proxies rather than direct measurements of community-level resources or educational systems.

The survey description is supported by the 2022 GDHS Final Report [23] and the Ghana Statistical Service microdata catalogue. The current global breastfeeding estimate and target are reported in the WHO global nutrition targets 2030 breastfeeding brief [24].

## 3. Results

### 3.1 Selection of the analytical sample

The Birth Recode dataset comprised 34,663 birth records [23]. Restriction to each respondent’s youngest living child, aged 0–5 completed months, yielded 959 mother–infant pairs. Because one infant was not living with the interviewed mother, that infant was excluded. Among the remaining 958 mothers, 620 reported that they were currently working and constituted the sample for the prevalence, descriptive, and bivariate analyses. One mother had an indeterminate response for the number of antenatal care visits. The complete-case multivariable analysis therefore included 619 mothers (Figure 1).

**Figure 1.**
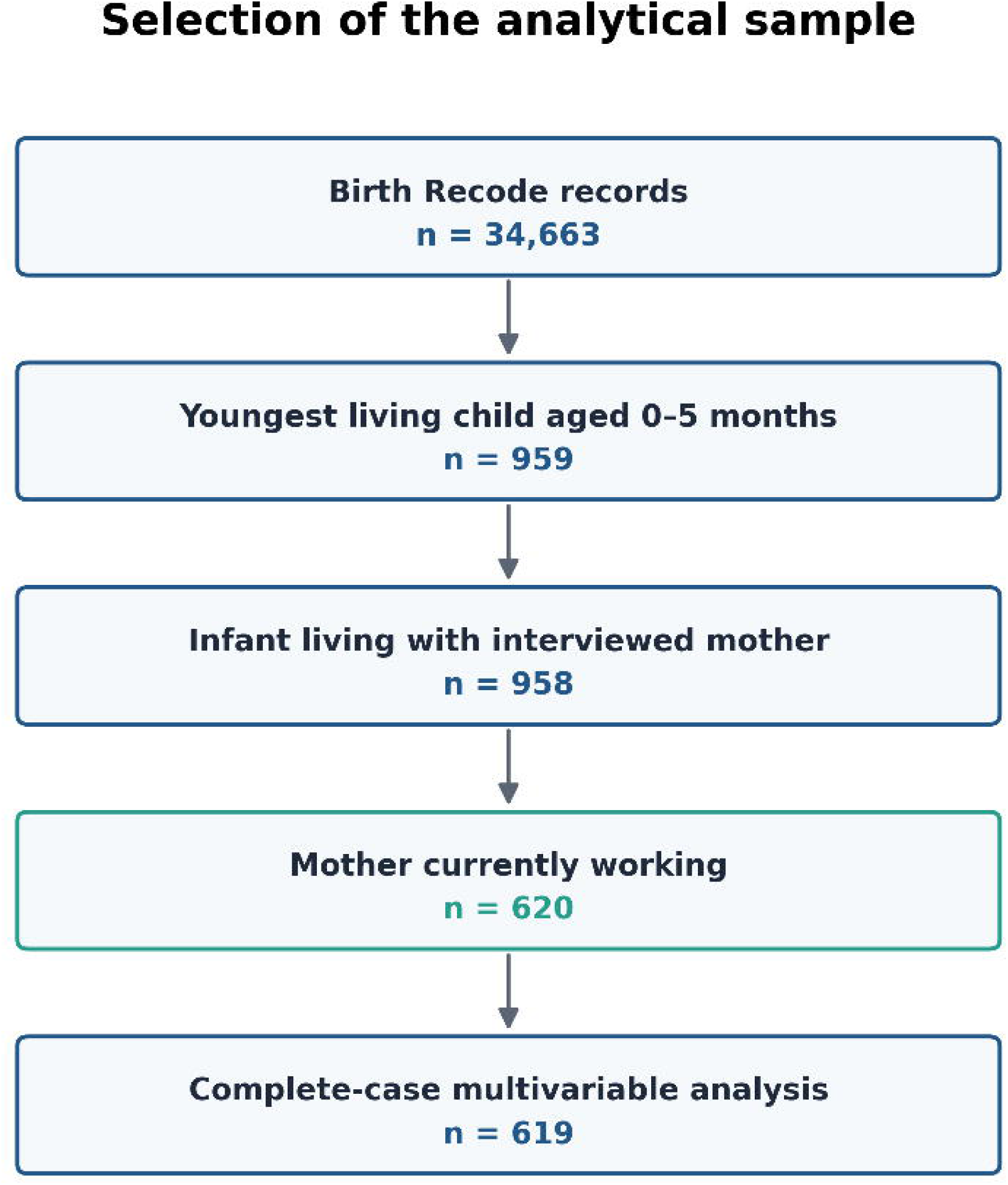
Selection of working mothers of infants aged 0–5 months from the 2022 Ghana Demographic and Health Survey.

#### Characteristics of the study participants

Most mothers were aged 20–34 years at the birth of the index child (70.4%). Nearly two-thirds had secondary or higher education (64.2%), while 20.7% had no formal education. Christianity appeared to be the predominant religion (67.3%), followed by Islam (27.5%). More than half of the mothers were multiparous (54.3%), while 23.4% were grand multiparous and 22.3% were primiparous.

Akan mothers comprised the largest ethnic group (37.3%), followed by Mole-Dagbani mothers (22.6%). Poor households accounted for 43.4% of the weighted sample, while 39.0% were classified as rich. Most mothers delivered vaginally (83.1%), and 56.1% of the index infants were male.

The findings suggest that 86.9% of mothers attended at least four antenatal care visits, while 52.6% reported receipt of postnatal care for their infants. Rural residents constituted 54.7% of the weighted sample; more than half resided in high-poverty communities (55.9%) and communities classified as having low educational attainment (55.4%) (Table 1).

**Table 1.**
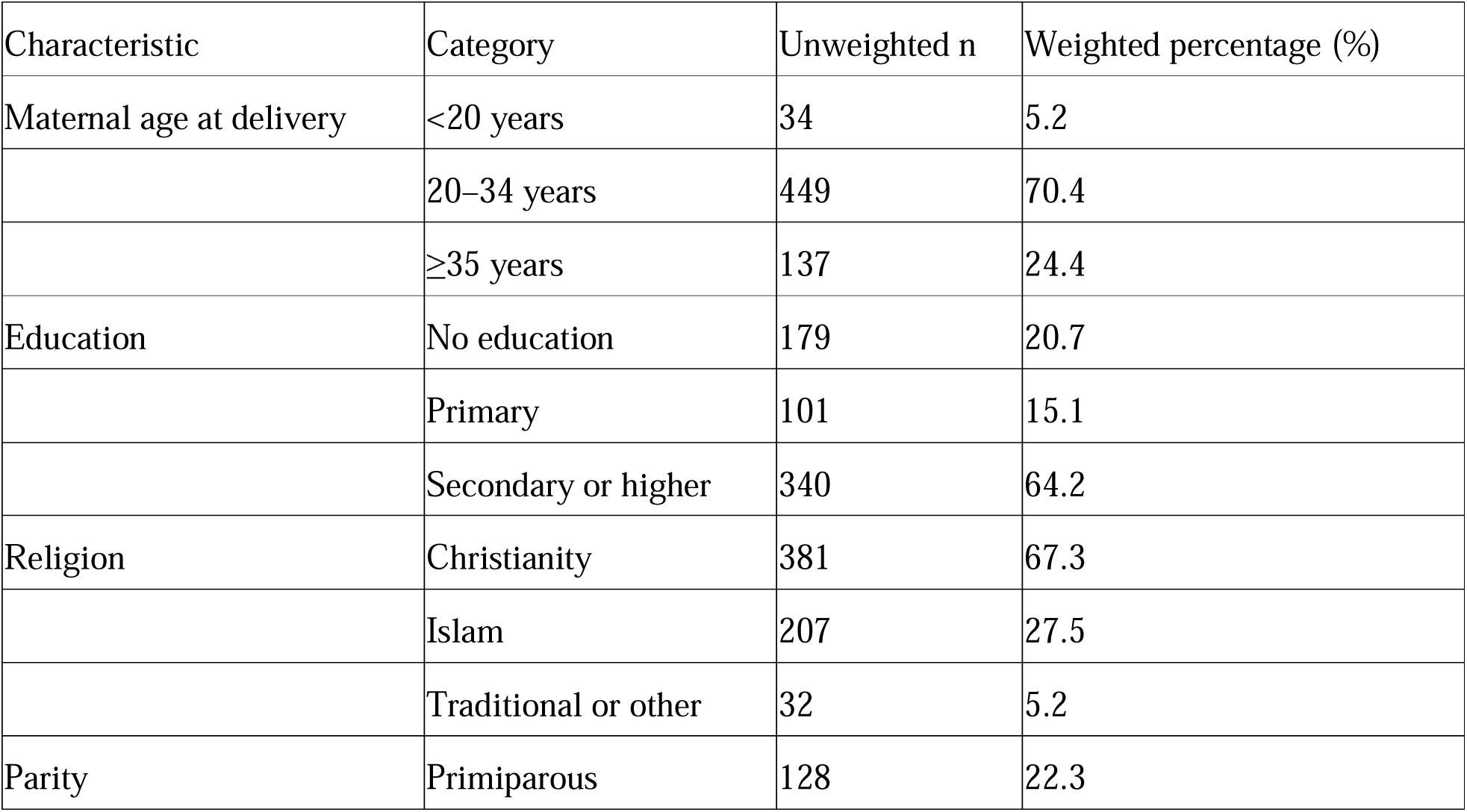
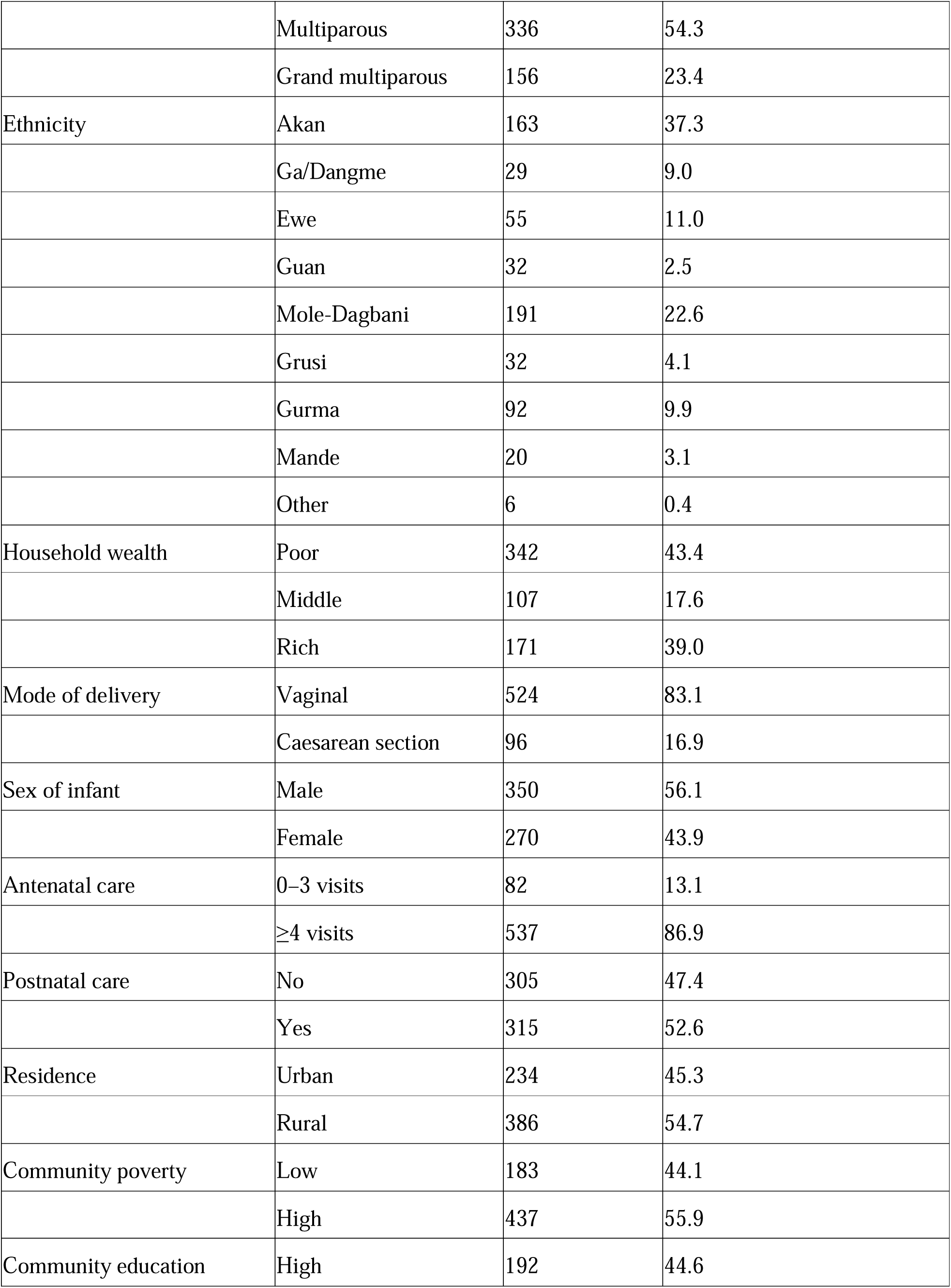
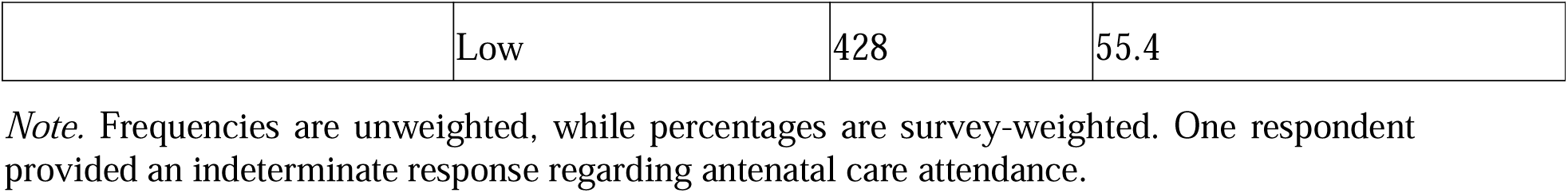
Characteristics of working mothers of infants aged zero–five months in Ghana.

#### Prevalence of exclusive breastfeeding

Of the 620 working mothers, 354 exclusively breastfed their infants during the 24 hours preceding the survey, while 266 did not. The survey-weighted prevalence of exclusive breastfeeding was estimated at 54.3% (95% confidence interval: 49.2–59.3) (Figure 2).

**Figure 2.**
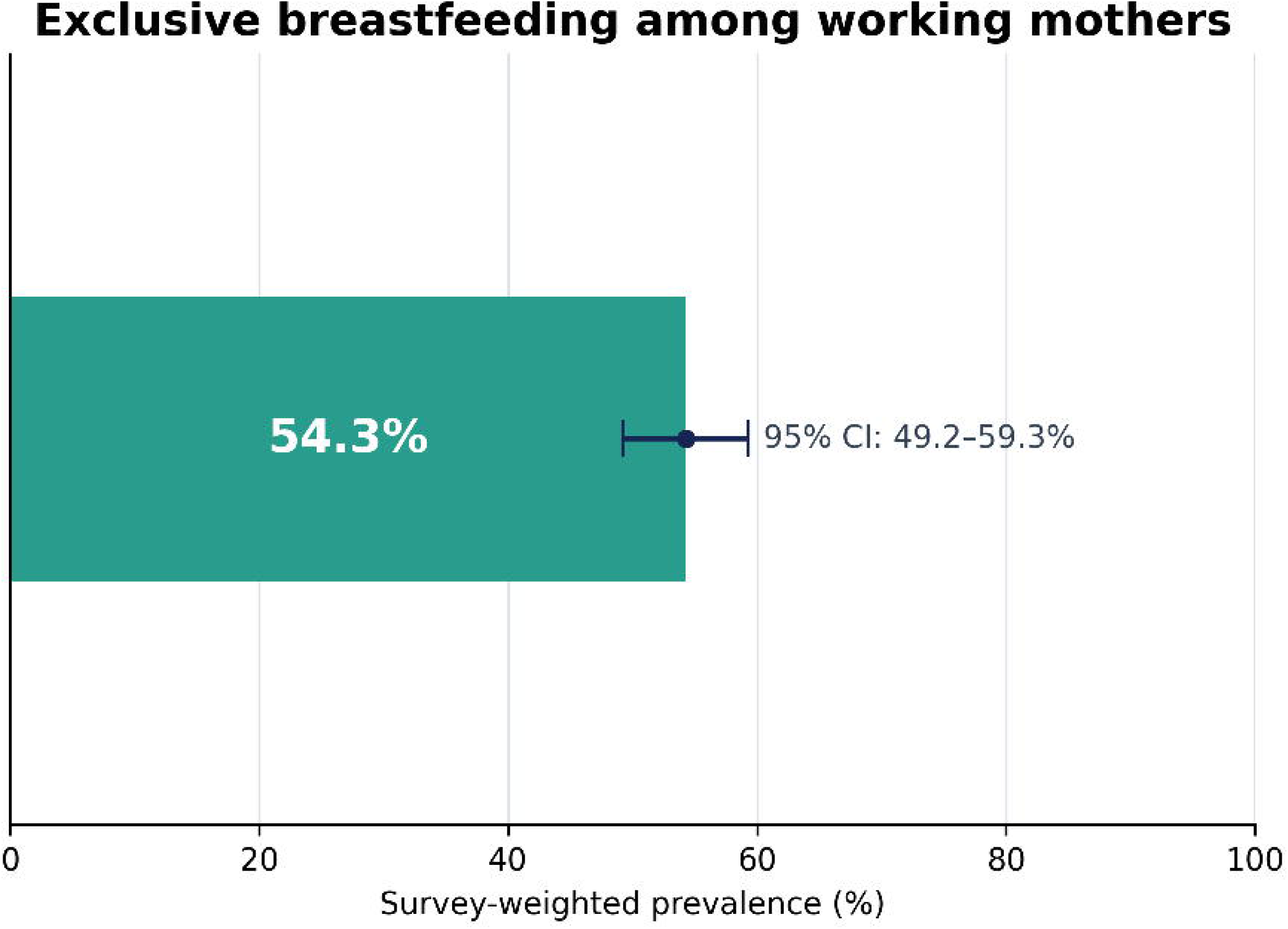
Survey-weighted prevalence of exclusive breastfeeding among working mothers of infants aged 0–5 months in Ghana (n = 620). The error bar presents the 95% confidence interval.

Exclusive breastfeeding prevalence appeared to increase across maternal age categories, from 43.6% among mothers younger than 20 years to 53.0% among those aged 20–34 years and 60.3% among those aged at least 35 years. The prevalence was 58.6% among mothers with no formal education, 51.5% among those with primary education, and 53.5% among those with secondary or higher education.

The prevalence was estimated at 62.9% among grand multiparous mothers, compared with 54.0% among multiparous mothers and 45.8% among primiparous mothers. Mothers from poor households appeared to have a higher prevalence than those from middle-wealth and rich households, at 58.6%, 48.4%, and 52.1%, respectively.

Prevalence appeared to vary substantially across ethnic and regional categories. Exclusive breastfeeding prevalence was highest among Guan mothers (91.6%), followed by Gurma (75.1%) and Mande mothers (75.0%). The estimates for these groups should be interpreted cautiously because of their relatively small, unweighted numbers.

Regional prevalence ranged from 25.8% in the Western North Region to 78.2% in the Oti Region. The Northern (77.3%), Savannah (73.0%), and Volta (72.6%) regions also recorded comparatively high prevalence. Western North, Greater Accra, Central, and Western exhibited prevalences below 45% (Figure 3).

**Figure 3.**
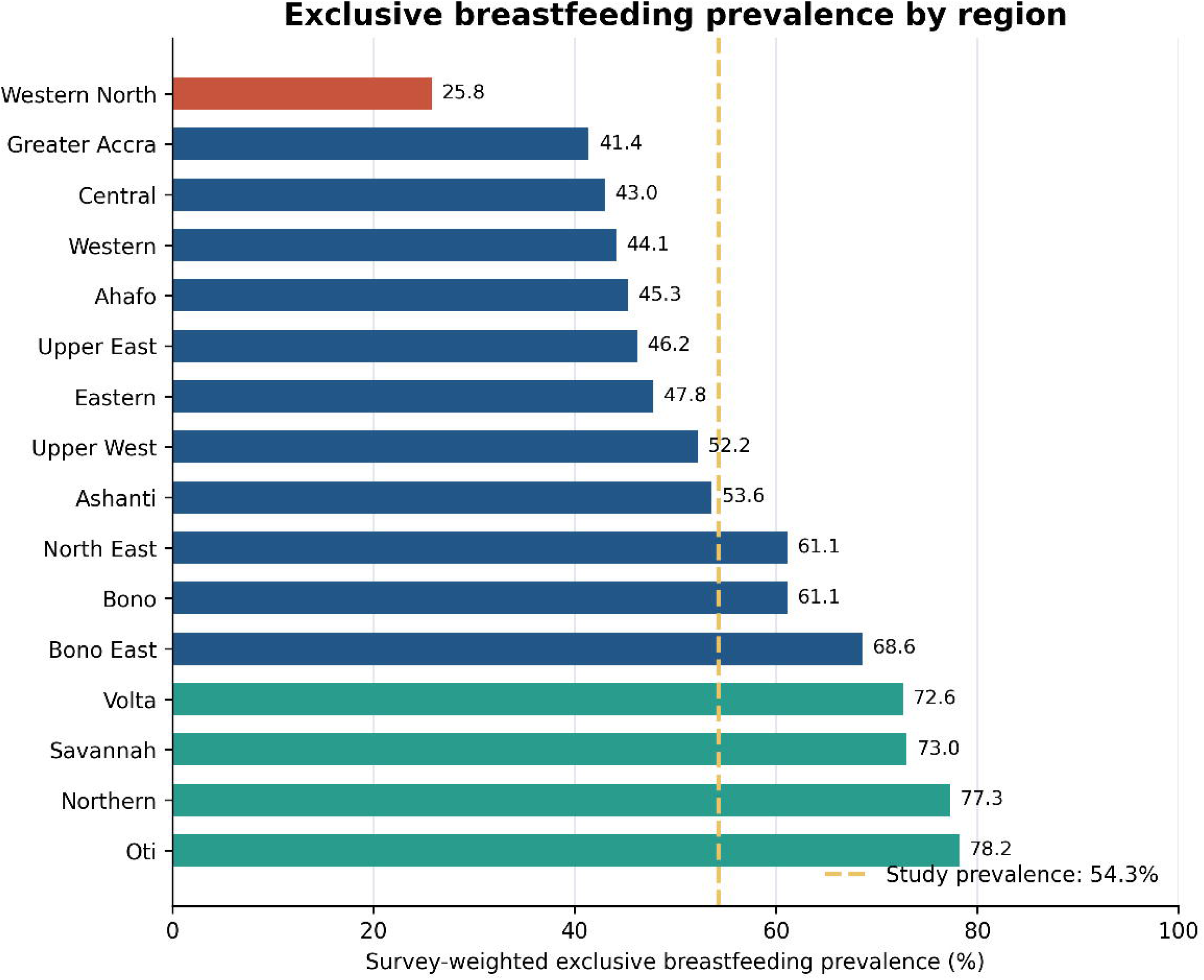
Survey-weighted prevalence of exclusive breastfeeding among working mothers by region of residence. The dashed line indicates the overall study prevalence of 54.3%.

Mothers who delivered vaginally recorded a higher prevalence than those who delivered by caesarean section, at 57.2% and 40.0%, respectively. Exclusive breastfeeding prevalence was 55.2% among mothers who attended at least four antenatal care visits and 48.0% among those who attended 0–3 visits. Little difference appeared to be observed according to postnatal care or place of residence.

#### Bivariate factors associated with exclusive breastfeeding

The design-adjusted bivariate analysis appeared to delineate significant associations between exclusive breastfeeding and ethnicity (Wald χ² = 29.98, df = 8, p < 0.001), mode of delivery (Wald χ² = 5.08, df = 1, p = 0.024), region of residence (Wald χ² = 51.18, df = 15, p < 0.001), and community poverty (Wald χ² = 4.06, df = 1, p = 0.044).

Parity demonstrated a possible association with exclusive breastfeeding, although the result did not reach the 5% threshold for statistical significance (Wald χ² = 5.12, df = 2, p = 0.077). Maternal age, education, religion, household wealth, infant sex, antenatal care, postnatal care, urban or rural residence, and community education did not appear to be significantly associated with exclusive breastfeeding in the bivariate analysis (Table 2).

**Table 2.**
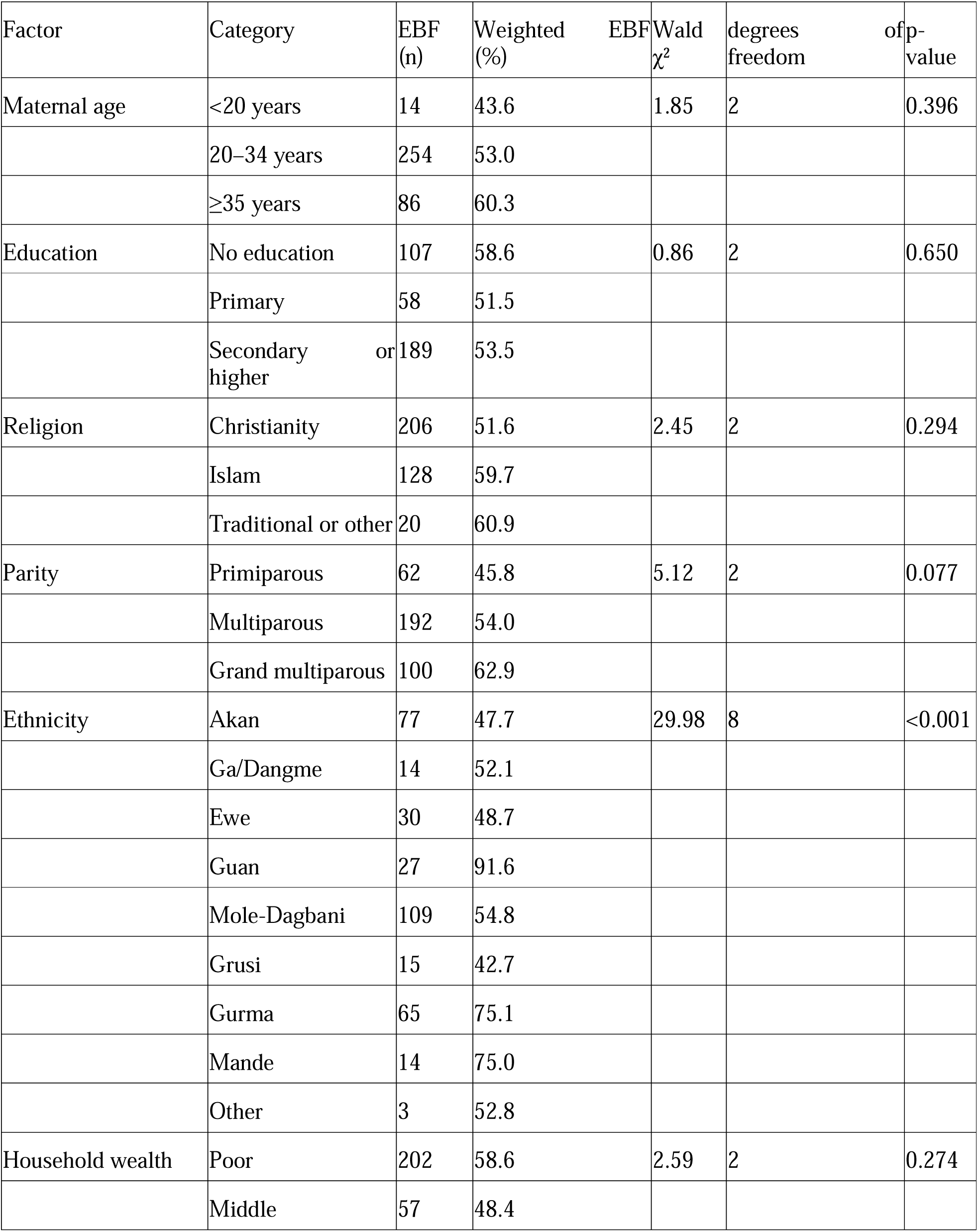
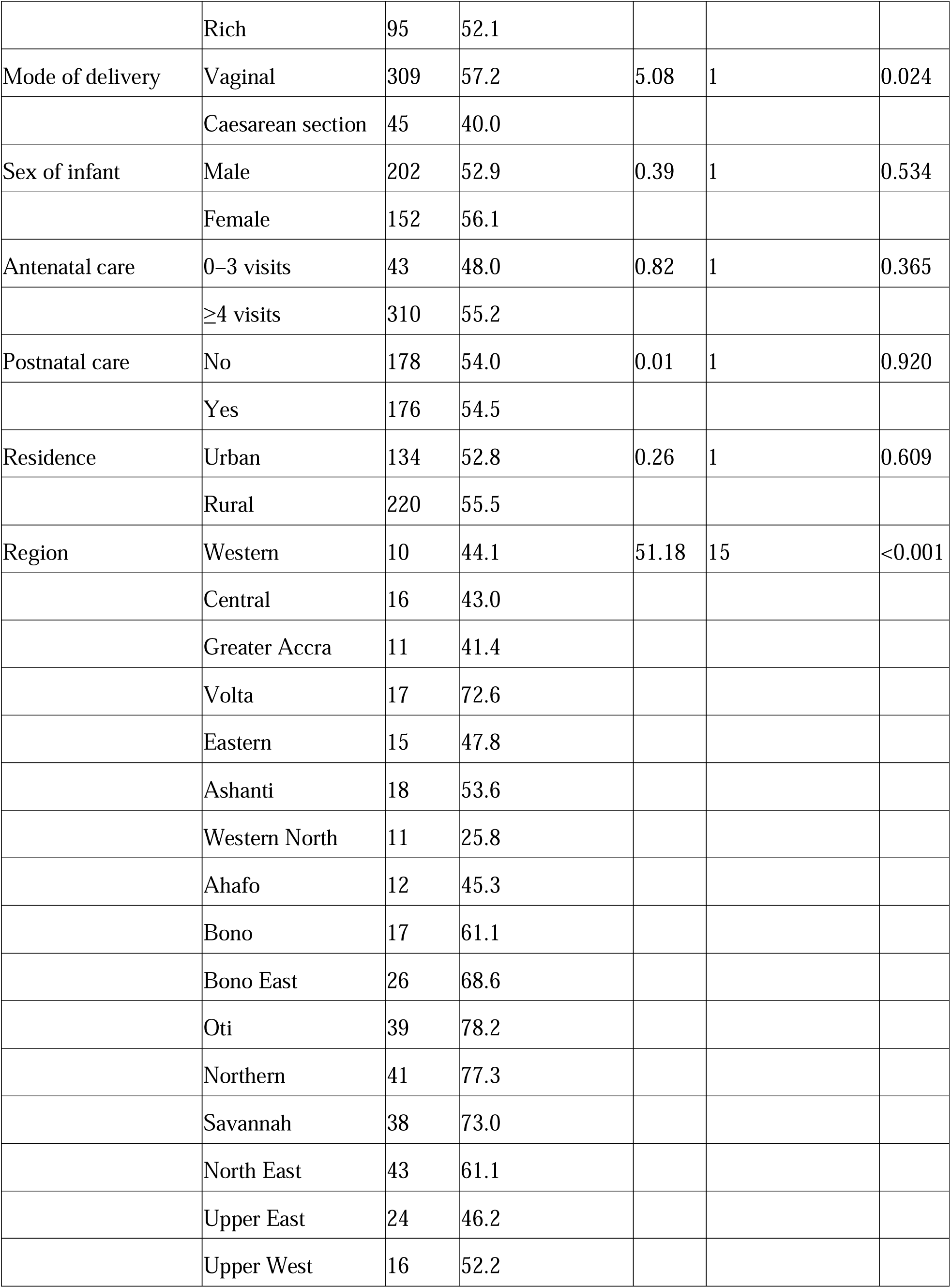
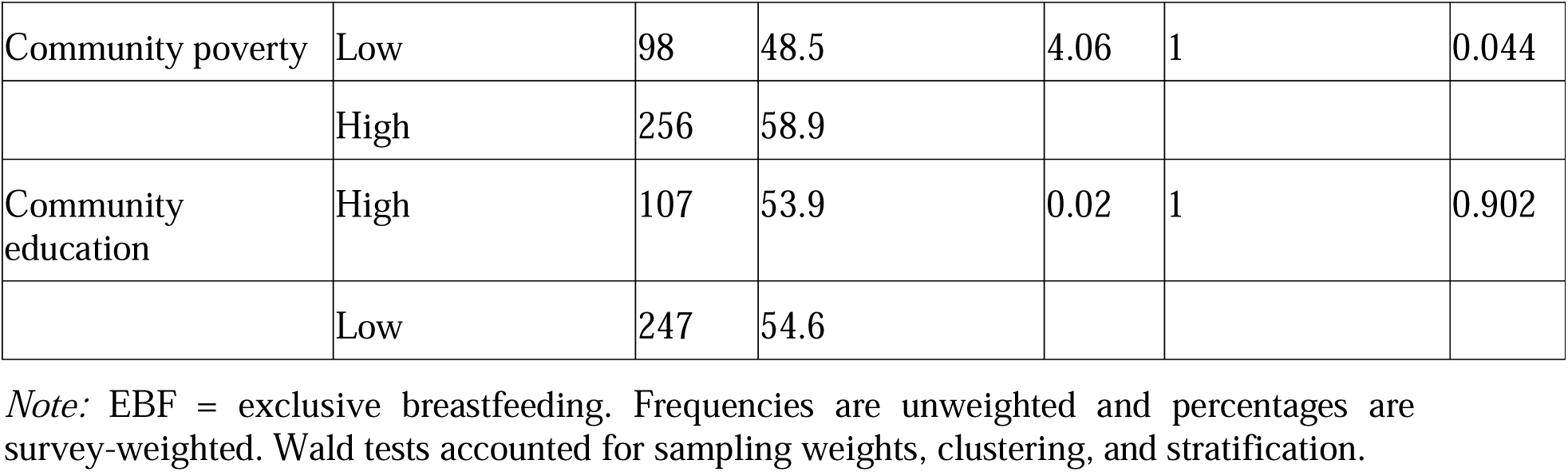
Bivariate associations between exclusive breastfeeding and employment among mothers.

#### Multivariable factors associated with exclusive breastfeeding

The complete-case multivariable analysis comprised 619 mothers; the overall model appeared to be statistically significant (Wald χ² = 82.34, df = 40, p < 0.001). The model produced a pseudo-Nagelkerke R² of 0.185 and correctly classified 67.1% of observations.

Region of residence appeared to remain jointly associated with exclusive breastfeeding after adjustment for the other individual, healthcare-related, and contextual variables (Wald χ² = 33.39, df = 15, p = 0.004). Mothers living in the Northern Region had approximately five times the odds of exclusive breastfeeding compared with mothers in the Western Region (AOR = 4.93; 95% CI: 1.50–16.17; p = 0.009). Mothers in the Savannah Region also had higher odds than those in the Western Region (AOR = 4.22; 95% CI: 1.08–16.41; p = 0.038).

The estimates for the Bono East and Oti regions suggested higher odds relative to the Western Region, but their confidence intervals included the null value. Consequently, neither association appeared to meet the 5% threshold for statistical significance.

Community education appeared to be independently associated with exclusive breastfeeding (p = 0.044). Mothers living in communities classified as having low educational attainment had 46.1% lower odds of exclusive breastfeeding than mothers living in high-education communities (AOR = 0.54; 95% CI: 0.30–0.98).

Guan mothers were estimated to have higher odds of exclusive breastfeeding than Akan mothers (AOR = 5.88; 95% CI: 1.38–25.11; p = 0.017). Nevertheless, ethnicity was not statistically significant when assessed as an overall variable in the adjusted model (Wald χ² = 12.05, df = 8, p = 0.149). The estimate for Guan mothers was based on 32 observations and had a wide confidence interval. It should therefore be interpreted cautiously.

Maternal age, education, religion, parity, household wealth, infant sex, antenatal care, postnatal care, urban or rural residence, and community poverty did not appear to be independently associated with exclusive breastfeeding. Caesarean delivery appeared to be associated with lower odds of exclusive breastfeeding, although the adjusted estimate did not meet the threshold for statistical significance (AOR = 0.57; 95% CI: 0.31–1.05; p = 0.071) (Table 3 and Figure 4).

**Figure 4.**
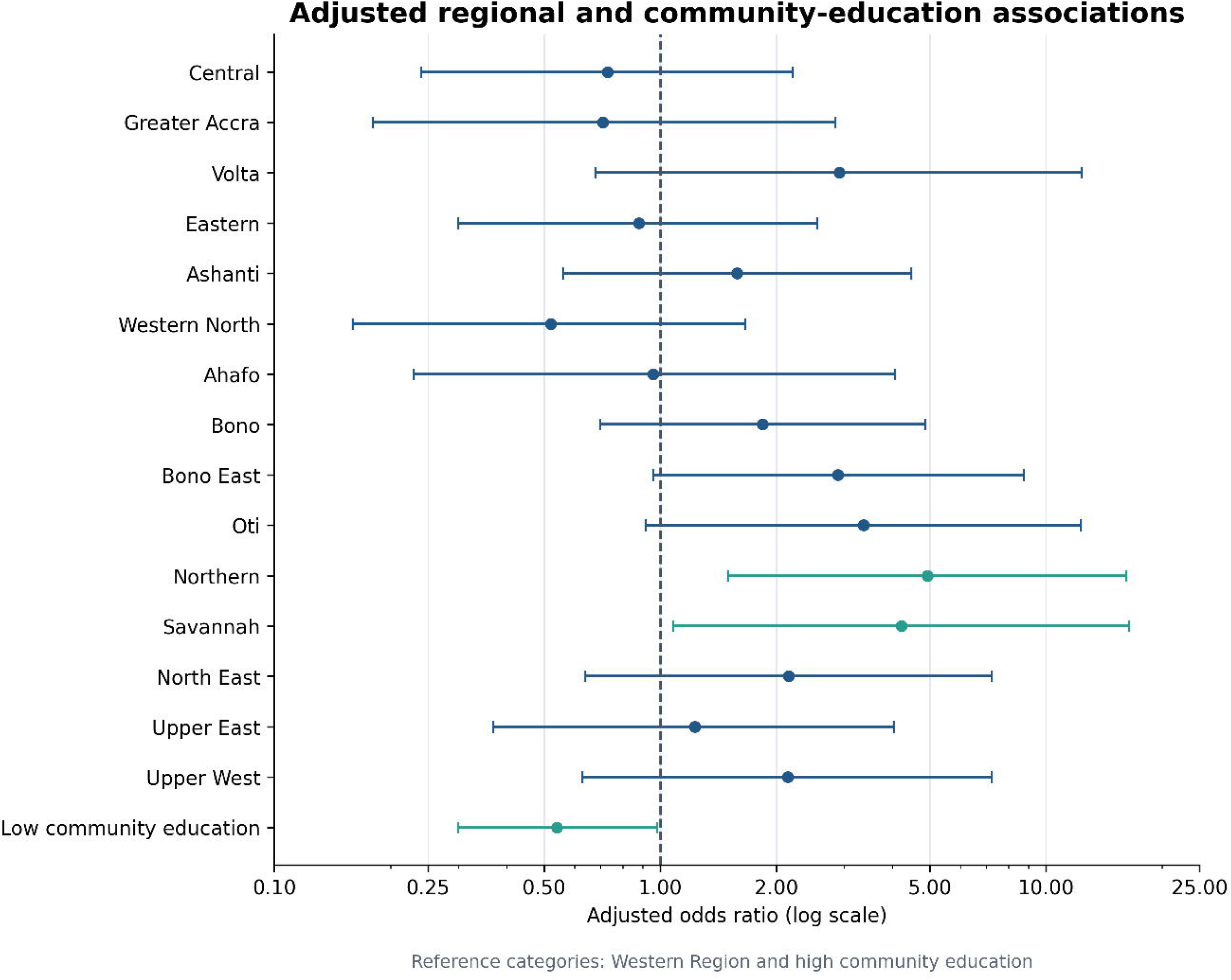
Adjusted regional and community-education associations with exclusive breastfeeding among working mothers. Points represent adjusted odds ratios, whereas horizontal lines denote 95% confidence intervals. Western Region and high community education were the reference categories. The vertical dashed line indicates no association (AOR = 1.00).

**Table 3.** Survey-weighted logistic regression of factors associated with exclusive breastfeeding.

| Characteristic | Category | Adjusted odds ratio (AOR) | 95% confidence interval (CI) | p-value |
| --- | --- | --- | --- | --- |
| Maternal age | <20 years | 1.00 | Reference |  |
|  | 20–34 years | 1.43 | 0.49–4.16 | 0.513 |
|  | ≥35 years | 1.82 | 0.48–6.90 | 0.377 |
| Education | No education | 1.00 | Reference |  |
|  | Primary | 1.52 | 0.68–3.41 | 0.312 |
|  | Secondary or higher | 1.64 | 0.83–3.25 | 0.152 |
| Religion | Christianity | 1.00 | Reference |  |
|  | Islam | 0.98 | 0.50–1.93 | 0.960 |
|  | Traditional or other | 0.66 | 0.23–1.91 | 0.444 |
| Parity | Primiparous | 1.00 | Reference |  |
|  | Multiparous | 1.52 | 0.81–2.84 | 0.193 |
|  | Grand multiparous | 2.10 | 0.91–4.82 | 0.082 |
| Ethnicity | Akan | 1.00 | Reference |  |
|  | Ga/Dangme | 1.57 | 0.48–5.15 | 0.457 |
|  | Ewe | 1.07 | 0.39–2.96 | 0.889 |
|  | Guan | 5.88 | 1.38–25.11 | 0.017 |
|  | Mole-Dagbani | 1.00 | 0.45–2.23 | 1.000 |
|  | Grusi | 0.72 | 0.23–2.28 | 0.573 |
|  | Gurma | 2.71 | 0.90–8.16 | 0.076 |
|  | Mande | 3.31 | 0.65–16.80 | 0.149 |
|  | Other | 1.02 | 0.08–13.16 | 0.986 |
| Household wealth | Poor | 1.00 | Reference |  |
|  | Middle | 0.87 | 0.41–1.85 | 0.708 |
|  | Rich | 1.24 | 0.53–2.92 | 0.620 |
| Mode of delivery | Vaginal | 1.00 | Reference |  |
|  | Caesarean section | 0.57 | 0.31–1.05 | 0.071 |
| Sex of infant | Male | 1.00 | Reference |  |
|  | Female | 1.12 | 0.71–1.76 | 0.627 |
| Antenatal care | 0–3 visits | 1.00 | Reference |  |
|  | ≥4 visits | 1.55 | 0.77–3.11 | 0.220 |
| Postnatal care | No | 1.00 | Reference |  |
|  | Yes | 1.13 | 0.72–1.78 | 0.592 |
| Residence | Urban | 1.00 | Reference |  |
|  | Rural | 0.84 | 0.44–1.60 | 0.598 |
| Region | Western | 1.00 | Reference |  |
|  | Central | 0.73 | 0.24–2.21 | 0.573 |
|  | Greater Accra | 0.71 | 0.18–2.85 | 0.631 |
|  | Volta | 2.91 | 0.68–12.34 | 0.148 |
|  | Eastern | 0.88 | 0.30–2.55 | 0.817 |
|  | Ashanti | 1.58 | 0.56–4.47 | 0.390 |
|  | Western North | 0.52 | 0.16–1.66 | 0.269 |
|  | Ahafo | 0.96 | 0.23–4.07 | 0.954 |
|  | Bono | 1.84 | 0.70–4.87 | 0.217 |
|  | Bono East | 2.89 | 0.96–8.75 | 0.060 |
|  | Oti | 3.36 | 0.92–12.32 | 0.068 |
|  | Northern | 4.93 | 1.50–16.17 | 0.009 |
|  | Savannah | 4.22 | 1.08–16.41 | 0.038 |
|  | North East | 2.15 | 0.64–7.23 | 0.217 |
|  | Upper East | 1.23 | 0.37–4.03 | 0.737 |
|  | Upper West | 2.14 | 0.63–7.24 | 0.222 |
| Community poverty | Low | 1.00 | Reference |  |
|  | High | 1.71 | 0.81–3.59 | 0.160 |
| Community education | High | 1.00 | Reference |  |
|  | Low | 0.54 | 0.30–0.98 | 0.044 |

*Note.* AOR = adjusted odds ratio; CI = confidence interval. The model accounted for sampling weights, primary sampling units, and sampling strata. Reference categories are presented as AOR = 1.00.

## 4. Conclusion

This study found that 54.3% of working Ghanaian mothers with infants aged 0–5 months practised exclusive breastfeeding, a prevalence close to the 2022 national estimate but below the 60% global target for 2030 [9]. Regional and community conditions appeared to predominate over most measured individual characteristics. Mothers in the Northern and Savannah regions had higher adjusted odds of exclusive breastfeeding than those in the Western Region, while mothers in communities with low educational attainment had lower odds than those in high-education communities. The higher estimate among Guan mothers should be interpreted cautiously because ethnicity was not significant overall and the subgroup was small. Maternal age, education, religion, parity, household wealth, infant sex, antenatal care, postnatal care, and residence were not independently associated with exclusive breastfeeding. Caesarean delivery was associated only in the unadjusted analysis.

Findings appear to substantiate the case for regionally responsive breastfeeding programmes, community-level educational initiatives, enhanced counselling provision, and workplace measures such as breastfeeding breaks, flexible return-to-work arrangements and facilities for expressing and storing milk. These workplace recommendations are informed by previous evidence [25] because employment conditions were not measured directly. Future studies could profitably examine occupation, maternity leave, work schedules, childcare, family support, and workplace facilities using longitudinal and qualitative methodologies. Cross-sectional findings require cautious interpretation.

## Data Availability

A quantitative, cross-sectional analytical study was undertaken using secondary data from the 2022 Ghana Demographic and Health Survey.

https://dhsprogram.com

## Author Contributions

Abubakar Iddrisu Siddiq conceptualised the study, developed the methodology, analysed and interpreted the data, and prepared the original manuscript. Ishmael Saafu contributed to the study design, data analysis, interpretation of the findings, and critical revision of the manuscript. Edmund Teye Borkor, Emmanuel Vondee, Felicia Tenpoka Sampana, and Bernice Okine contributed to the literature review, interpretation of the findings, and critical revision of the manuscript. All authors reviewed and approved the final manuscript and accept responsibility for the integrity of the work.

